# Replicating the Association of Variants in *BSN* and *APBA1* with Obesity in Diverse Populations

**DOI:** 10.1101/2024.08.21.24312322

**Authors:** Jamie R. Robinson, Joshua C. Denny, Chenjie Zeng

## Abstract

In a recent study by Zhao et al., rare protein-truncating variants (PTVs) in the BSN and APBA1 genes showed effects on obesity that exceeded those of well-known genes such as MC4R in a UK cohort. In this study, we leveraged the All of Us Research Program, to investigate the association of predicted LoF (pLoF) PTVs in BSN and APBA1 with body mass index (BMI) across a population of diverse ancestry. Our analysis revealed that the impact of pLoF variants in BSN and APBA1 on BMI was notably greater in this cohort, especially among individuals of European ancestry. Additionally, a phenome-wide association study (PheWAS) using the extensive phenotypic data available in the All of Us Research Program uncovered novel associations of *BSN* and *APBA1*heterozygous pLoF carriers with various phenotypes. Specifically, BSN pLoF variants were associated with pulmonary hypertension, atrial fibrillation, and anticoagulant use, while APBA1 pLoF variants were linked to disorders of the temporomandibular joint. These findings underscore the potential of large-scale biobanks in advancing genetic discovery.

## Introduction

Obesity is a highly heritable condition and major risk factor for many common diseases.^1^ In their recent publication examining protein-truncating variants (PTVs) in *BSN* and *APBA1*, Zhao et al. observed rare loss-of-function (LoF) variants in two genes (*BSN* and *APBA1*) with effects substantially larger than those of well-established obesity genes such as *MC4R* and replicated these findings in two non-European cohorts.^2^ Through analyses in the *All of Us* Research Program, we analyzed the association of predicted LOF (pLoF) PTVs in *BSN* and *APBA1* with body max index (BMI) to assess if the association was replicated in another population of diverse ancestry background. We show that the increase in BMI associated with pLoF variants in *BSN* and *APBA1* was considerably larger in this cohort, particularly in those of European ancestry. We also performed a phenome-wide association study (PheWAS), leveraging the rich phenotypic data within the *All of Us* Research Program, identifying novel phenotypic associations of *BSN* and *APBA1* heterozygous pLoF carriers. With this, we identified associations of *BSN* with pulmonary hypertension, atrial fibrillation, and use of anticoagulants, while *APBA1* pLoF variants were associated with disorders of the temporomandibular joint.

### Study Population

We utilized data from 206,173 adults of diverse ancestry in the *All of Us* Research Program, an ongoing population-based longitudinal cohort study conducted in the United States aiming to recruit at least 1 million participants reflecting the diversity of the population.^3,4^ A detailed description of the consent and enrollment process has been described elsewhere.^3^

### Whole-Genome Sequencing and Quality Control

The *All of Us* Genome Centers conducted whole-genome sequencing (WGS). Quality control (QC), mapping, and concordance with genotyping arrays to ensure accuracy has been described previously.^4^ These procedures ensured that the final WGS data maintained clinical-grade quality with consistent coverage (≥30x mean) and uniformity across sequencing centers. We removed variants with a read depth of less than 10x, genotype quality below 20, allele balance of 0.2 or lower for heterozygotes, Phred-scaled p-value for exact tests of excess heterozygosity below 54.69, QUAL score below 60 for single nucleotide polymorphisms and below 69 for insertions and deletions, a call rate below 0.99, and a minor allele frequency > 0.01.

### Genetic Similarity Assessment

We categorized individuals by genetic similarity to global population samples including 1000 Genomes Project and Human Genome Diversity Project as inferred by principal component analysis (PCA). Six groups were included: African (AFR), American/American Admixed (AMR), East Asian (EAS), European (EUR), and Middle East (MID) and Southeast Asian (SAS). This approach, while useful, does not equate to defining race or ethnicity and oversimplifies the complex nature of human genetic diversity.

### Variant annotation

Variant annotation was performed using the Ensembl Variant Effect Predictor (VEP version 110) with the plugin of the Loss-Of-Function Transcript Effect Estimator (LOFTEE). Each variant was mapped to a single Ensembl transcript. We prioritized transcripts that were part of the MANE Select transcripts or designated as the canonical transcripts by VEP.^5^ Variants predicted to be stop-gained, frame-shift, splice acceptor, and splice donor by LOFTEE were grouped into the predicted loss-of-function (pLoF) category.

### Statistical analysis

For BMI calculation, we used data of weight and height of participants collected at enrollment. To test whether there was an association between pLoF variants in the genes *BSN* and *APBA1* with BMI at enrollment, we performed a gene-based analysis by collapsing all pLoF variants for each gene. We tested if participants with pLoF variants had a higher BMI at enrollment compared to those without such variants by the Mann–Whitney U test. We also tested associations by linear regression model, adjusted for age at enrollment and self-reported sex at birth. We identified 800 and 723 phenotypes, respectively, for *BSN* and *APB1* carriers. For PheWAS of pLoF carriers, we used firth logistic regression, adjusting for age, self-reported sex at birth and the top 16 PCs. Owing to correlations among phenotypes assessed, we determined the empirical PheWAS significance threshold through simulations converging as 8.50 × 10^−5^. Statistical testing was one-sided for BMI and two-sided for PheWAS. Analyses were conducted within the *All of Us* workbench with Python (version 3.9.10) or R (version 4.2.0). We received an exception from the Resource Access Board to disseminate select participants counts less than 20.

## Results

Among the 202,000 individuals with available genomic and phenotypic data in the *All of Us* cohort, there were 67 and 61 individuals with a *BSN* or *APBA1* pLoF variant, respectively (Table 1). Of the entire cohort, mean BMI was 3.6 kg m^−2^ and 3.2 kg m^−2^ higher than non-carriers for *BSN* and *APBA1* heterozygous carriers, respectively (Figure 1A & 1B), findings which were more pronounced compared to the UK Biobank (mean BMI increase of 3.1 and 2.1 kg m^−2^, respectively for *BSN* and *APBA1*). Associations of BMI were driven by findings in those of predominantly European genetic ancestry. We found that the direction of associations persisted in those of non-predominantly European genetic ancestry for *BSN* but not for *APBA1* (Figure 1C & D).

**Table 1.** Ancestry and Demographic Characteristics of Cohort. Genetic ancestry is based on similarity to reference populations of the 1000 Genomes and Human Genome Diversity Project.

| **Characteristics** | ***BSN*** |  | ***APBA1*** |  |
| --- | --- | --- | --- | --- |
|  | pLoF<br>(n = 67) | No pLoF<br>(n = 202,723) | pLoF<br>(n = 61) | No pLoF<br>(n = 205,641) |
| **Genetic ancestry** |  |  |  |  |
| African | 28 | 45,164 | 10 | 45,182 |
| Admixed Americans | 3 | 34,638 | 7 | 34,634 |
| East Asian | 1 | 4,123 | 0 | 4,124 |
| European | 35 | 113,687 | 44 | 113,678 |
| Middle East | 0 | 786 | 0 | 786 |
| Southeast Asian | 0 | 2,509 | 0 | 2,509 |
| **Age at enrollment yr, mean (SD)** | 50.6 (15.1) | 51.9 (16.9) | 46.8 (16.9) | 51.9 (16.9) |
| **Self-reported sex at birth** |  |  |  |  |
| Female | 37 | 120,615 | 40 | 120,612 |
| Male | 27 | 76,160 | 20 | 76,167 |

**Figure 1.**
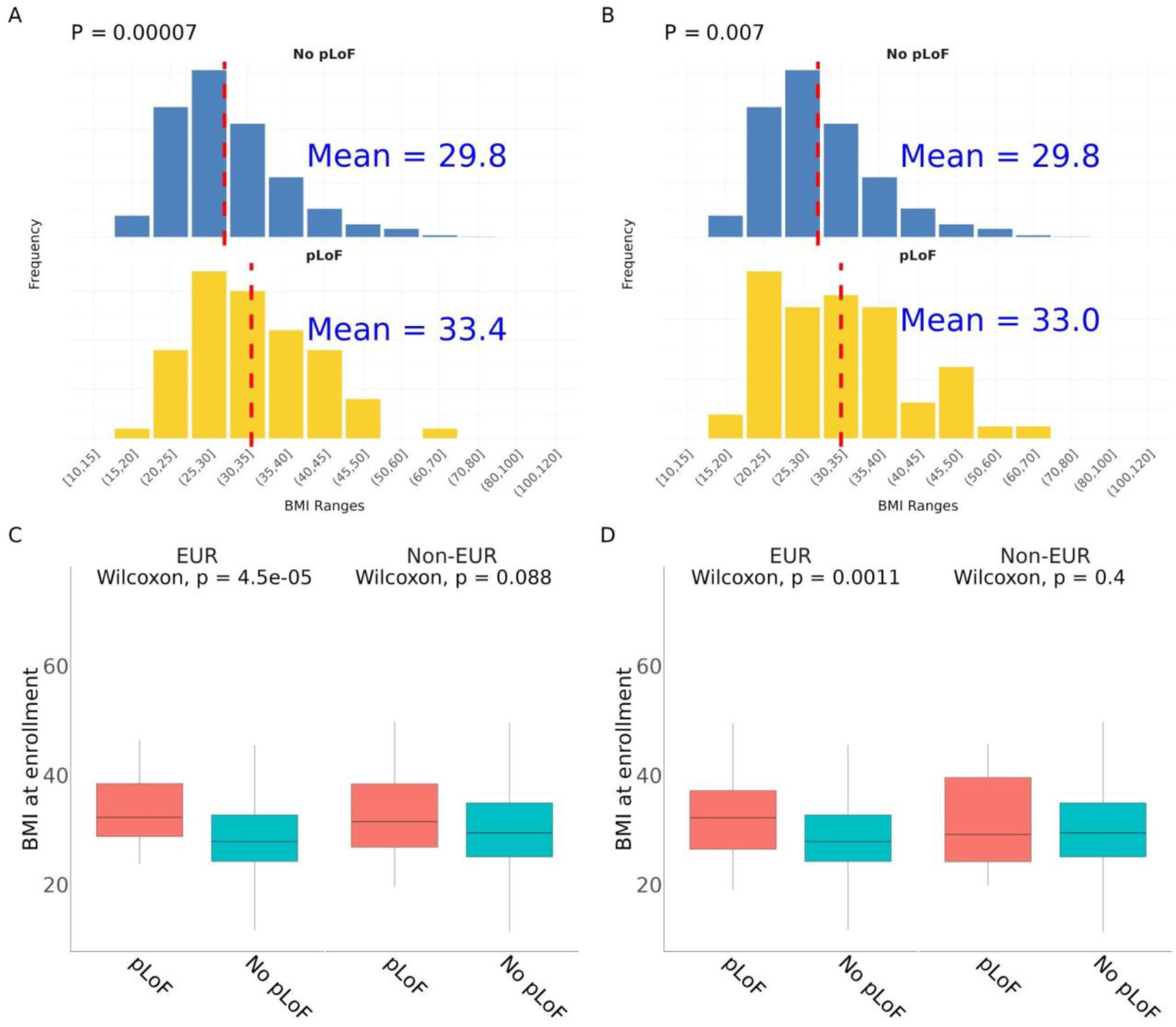
Association of predicted loss of function (pLoF) variants in *BSN* (A, C) and *APBA1* (B, D) with body mass index. Panels A and B use the entire *All of Us cohort* and panels C and D compare body mass index in populations of predominantly European genetic ancestry to populations of non-predominantly European genetic ancestry, as based on similarity to reference populations.

PheWAS in pLoF carriers identified potentially new associations with these two genes (Figure 2). Associations exceeding phenome-wide significance threshold with *BSN* included secondary pulmonary arterial hypertension (OR 24.8, 95% CI: [6.8-63.8], *p*=8.2.×10^−5^), use of anticoagulants (OR 4.9 [2.5-8.7], *p*=1.6×10^−5^), and atrial fibrillation (OR 4.8, 95% CI: [2.4-9.1], *p*=4.6×10^−5^). A novel phenotype association of *APBA1* pLoFs was adhesions and ankylosis of the temporomandibular joint (OR 134.9, 95% CI: [15.0-680.6], *p*=1.0×10^−5^). Associations with *BSN* or *APBA*1 pLoF carriers remained statistically significant after conditioning on the BMI.

**Figure 2.**
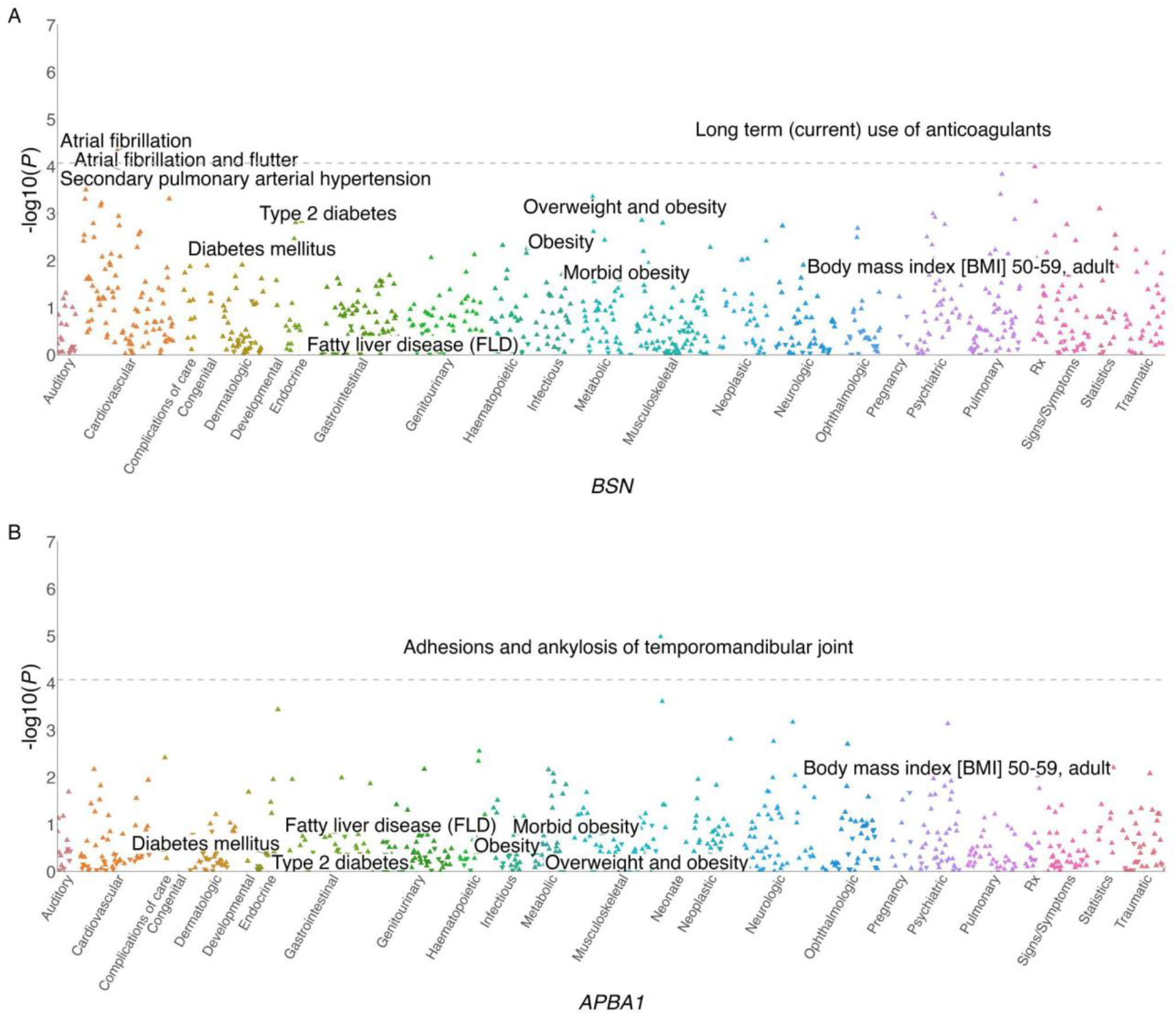
Phenome-wide association study (PheWAS) of predicted loss of function (pLoF) variants in *BSN* (A) and *APBA1* (B). Analysis represents Firth logistic regression adjusted for age, sex at birth and top 16 principal components. Dashed line represents the empirical significance threshold for PheWAS.

## Discussion

We replicated findings by Zhao *et al*. demonstrating *BSN* and *APBA1* to be associated with a substantial increase in BMI in the *All of Us* cohort, with effect size of the association greater than those in the well-studied components of the leptin-melanocortin pathway^6–8^ and a slightly larger effect size in *All of Us* than those reported in Zhao et al.^2^

Variants in *BSN* were found to be associated with atrial fibrillation and pulmonary artery hypertension, potential comorbidities of obesity. Interestingly, associations remained after adjusting for BMI, suggesting possible pleiotropic effects. *BSN*, encoding the bassoon protein involved in neurotransmitter release and autonomic regulation may influence cardiac rhythm and pulmonary vascular remodeling leading to these phenotypes.^9^ Variants in *APBA1*, in contrast, were associated with disorders of the temporomandibular joint, not typically considered an obesity comorbidity. *APBA1* encodes a neuronal adaptor protein involved in neuroprotection and synaptic function that inhibits production of proteolytic amyloid precursor protein fragments.^10^ This may result in an impact on neuronal health in the trigeminal system or tissue maintenance and inflammatory pathways potentially contributing to temporomandibular joint ankylosis. Further research would be needed to explore these potential connections.

The main limitation of this study is the small sample size of carriers, particularly in populations of non-predominantly European genetic ancestry. The non-European population in *All of Us* consists of a larger proportion of AFR genetic ancestry than Zhao et al., possibly contributing to disparate findings. As *All of Us* increases cohort size and other diverse populations become available, further studies are needed in additional ancestry populations.

In conclusion, the findings of associations with pLoFs in *BSN* and *APBA1* and BMI in the *All of Us* cohort lends further support to the importance of these genes in adiposity and identifies potentially new phenotype associations.

## Data Availability

All data produced in the present study are available upon reasonable request to the authors

## Acknowledgements

This research was supported by the Intramural Research Program of the National Human Genome Research Institute (NHGRI) and National Institutes of Health ZIA HG200417.

## References

1. GBD 2015 Obesity Collaborators et al. Health Effects of Overweight and Obesity in 195 Countries over 25 Years. N. Engl. J. Med. 377, 13–27 (2017).

2. Zhao, Y. et al. Protein-truncating variants in BSN are associated with severe adult-onset obesity, type 2 diabetes and fatty liver disease. Nat. Genet. 56, 579–584 (2024).

3. All of Us Research Program Investigators et al. The ‘All of Us’ Research Program. N. Engl. J. Med. 381, 668–676 (2019).

4. All of Us Research Program Genomics Investigators. Genomic data in the All of Us Research Program. Nature 627, 340–346 (2024).

5. Morales, J. et al. A joint NCBI and EMBL-EBI transcript set for clinical genomics and research. Nature 604, 310–315 (2022).

6. Vaisse, C., Clement, K., Guy-Grand, B. & Froguel, P. A frameshift mutation in human MC4R is associated with a dominant form of obesity. Nat. Genet. 20, 113–114 (1998).

7. Yeo, G. S. et al. A frameshift mutation in MC4R associated with dominantly inherited human obesity. Nat. Genet. 20, 111–112 (1998).

8. Chermon, D. & Birk, R. Predisposition of the Common MC4R rs17782313 Female Carriers to Elevated Obesity and Interaction with Eating Habits. Genes 14, 1996 (2023).

9. Hashida, H. et al. Cloning and mapping of ZNF231, a novel brain-specific gene encoding neuronal double zinc finger protein whose expression is enhanced in a neurodegenerative disorder, multiple system atrophy (MSA). Genomics 54, 50–58 (1998).

10. Ho, A., Liu, X. & Südhof, T. C. Deletion of Mint Proteins Decreases Amyloid Production in Transgenic Mouse Models of Alzheimer’s Disease. J. Neurosci. 28, 14392–14400 (2008).

